# An Anthropomorphic Diagnosis System of Pulmonary Nodules using Weak Annotation-Based Deep Learning

**DOI:** 10.1101/2024.05.03.24306828

**Authors:** Lipeng Xie, Yongrui Xu, Mingfeng Zheng, Yundi Chen, Min Sun, Michael A. Archer, Yuan Wan, Wenjun Mao, Yubing Tong

## Abstract

**Purpose:** To develop an anthropomorphic diagnosis system of pulmonary nodules (PN) based on Deep learning (DL) that is trained by weak annotation data and has comparable performance to full-annotation based diagnosis systems.

**Methods:** The proposed system uses deep learning (DL) models to classify PNs (benign vs. malignant) with weak annotations, which eliminates the need for time-consuming and labor-intensive manual annotations of PNs. Moreover, the PN classification networks, augmented with handcrafted shape features acquired through the ball-scale transform technique, demonstrate capability to differentiate PNs with diverse labels, including pure ground-glass opacities, part-solid nodules, and solid nodules.

**Results:** The experiments were conducted on two lung CT datasets: (1) public LIDC-IDRI dataset with 1,018 subjects, (2) In-house dataset with 2740 subjects. Through 5-fold cross-validation on two datasets, the system achieved the following results: (1) an Area Under Curve (AUC) of 0.938 for PN localization and an AUC of 0.912 for PN differential diagnosis on the LIDC-IDRI dataset of 814 testing cases, (2) an AUC of 0.943 for PN localization and an AUC of 0.815 for PN differential diagnosis on the in-house dataset of 822 testing cases. These results demonstrate comparable performance to full annotation-based diagnosis systems.

**Conclusions:** Our system can efficiently localize and differentially diagnose PNs even in resource-limited environments with good robustness across different grade and morphology sub-groups in the presence of deviations due to the size, shape, and texture of the nodule, indicating its potential for future clinical translation.

**Summary:** An anthropomorphic diagnosis system of pulmonary nodules (PN) based on deep learning and weak annotation was found to achieve comparable performance to full-annotation dataset-based diagnosis systems, significantly reducing the time and the cost associated with the annotation.

**Key Points:** A fully automatic system for the diagnosis of PN in CT scans using a suitable deep learning model and weak annotations was developed to achieve comparable performance (AUC = 0.938 for PN localization, AUC = 0.912 for PN differential diagnosis) with the full-annotation based deep learning models, reducing around 30%∼80% of annotation time for the experts.

The integration of the hand-crafted feature acquired from human experts (natural intelligence) into the deep learning networks and the fusion of the classification results of multi-scale networks can efficiently improve the PN classification performance across different diameters and sub-groups of the nodule.

## Introduction

Pulmonary nodules (PN) are opacities with a diameter of less than 30 mm in the lungs (1). PN can be classified based on their radiologic morphology into three subtypes: pure ground-glass opacity (GGO), part-solid nodules (PSN), and solid nodules (SN) (2). While some MPN exhibit characteristic radiological features, such as spiculated margins and inhomogeneous density, certain benign PN (BPN) closely resemble malignant PN (MPN). The misdiagnosis rate for MPN remains alarmingly high with a range of ∼30% to ∼70%, varying with the nodule’s diameter and subtype (3). There is a strong demand for a computer-aided diagnostic system that can automatically catch potential PNs present in CT scans, extract salient features for computing, and subsequently provide accurate diagnoses for MPN (4).

Depending on the feature extraction technique used, the PN classification systems can be categorized as by usage of either traditional models or deep learning (DL) models. Various traditional models have been developed based on imaging processing models, handcrafted features, and feature-based classifiers (5–7), which require manual delineation of nodules in CT images as the initial input for systems (8), suffering from a lack of automation and poor computing efficiency. In contrast, DL-based models (9–15) can automatically extract discriminative features that are highly correlated with the task of PN classification and exhibit high computational efficiency due to the efficient forward propagation algorithm and the parallel computing framework inherent to DL. However, the impressive performance of these DL-based models still relies on the availability of large amounts of high-quality and full-annotated data. Such annotations, which closely align with the nodule shape, necessitate labor-intensive manual labeling on each 2D slice of the nodule.

Moreover, the existing DL-based systems primarily focus on a singular classification task and lack the ability for hierarchical classification of grading nodules and subtypes. Therefore, these DL-based systems encounter limitations in their clinical translation.

The weak-annotated data, which is easier to handle and more user-friendly to radiologists, can be annotated on only a few 2D slices of the nodule. However, weak-annotated data has been rarely used to train DL-based PN detection models due to the absence of accurate location and size information for PN. The purpose of this study was to develop a DL-based PN classification system that leverages weak annotations for automated differential diagnosis of PN using axial CT scans, achieving comparable performance to full-annotation based diagnosis systems.

## Materials and Methods

### Data collection and Weak Annotation

In this study, we utilized two lung CT datasets to evaluate the performance of our method (Table 1):

(1) **LIDC-IDRI dataset** (16): Clinical chest CT scans from 1,018 patients were annotated by experienced radiologist . There were 2,632 nodule cases in the dataset and each nodule in scans was represented by the contour and bounding-box and labeled with a grade class (benign and malignant) according to the average malignancy score and a morphological subtype class (GGO, PSN, and SN) according to the average texture score.
(2) **In-house dataset**: Axial CT images (695×695×5-46) from 2,740 subjects were collected and weak-annotated. We utilized a straightforward, effective, and user-friendly annotation approach in which each nodule in the CT image was marked with a 2D red circle in the middle slice of the nodule, without strict delineating the exact boundary or bounding-box (tight rectangular in 2D space) of the nodule (Figure 1A).

**Table 1.** Summary of the datasets used in this study.

| Datasets | LIDC-IDRI |  |  |  |  |  |  |  |  | In-house |  |  |  |  |  |
| --- | --- | --- | --- | --- | --- | --- | --- | --- | --- | --- | --- | --- | --- | --- | --- |
| Labels | Benign |  |  | Malignant |  |  | Uncertain |  |  | Benign |  |  | Malignant |  |  |
|  | GGO | PSN | SN | GGO | PSN | SN | GGO | PSN | SN | GGO | PSN | SN | GGO | PSN | SN |
| Training set | 62 | 744 | 0 | 44 | 0 | 348 | 68 | 311 | 0 | 61 | 63 | 240 | 470 | 517 | 293 |
| Validation set | 10 | 124 | 0 | 7 | 0 | 58 | 11 | 52 | 0 | 10 | 11 | 40 | 78 | 86 | 49 |
| Testing set | 32 | 374 | 0 | 22 | 0 | 174 | 35 | 156 | 0 | 30 | 31 | 120 | 235 | 259 | 147 |
| Total | 104 | 1242 | 0 | 73 | 0 | 580 | 114 | 519 | 0 | 101 | 105 | 400 | 783 | 862 | 489 |

**Figure 1:**
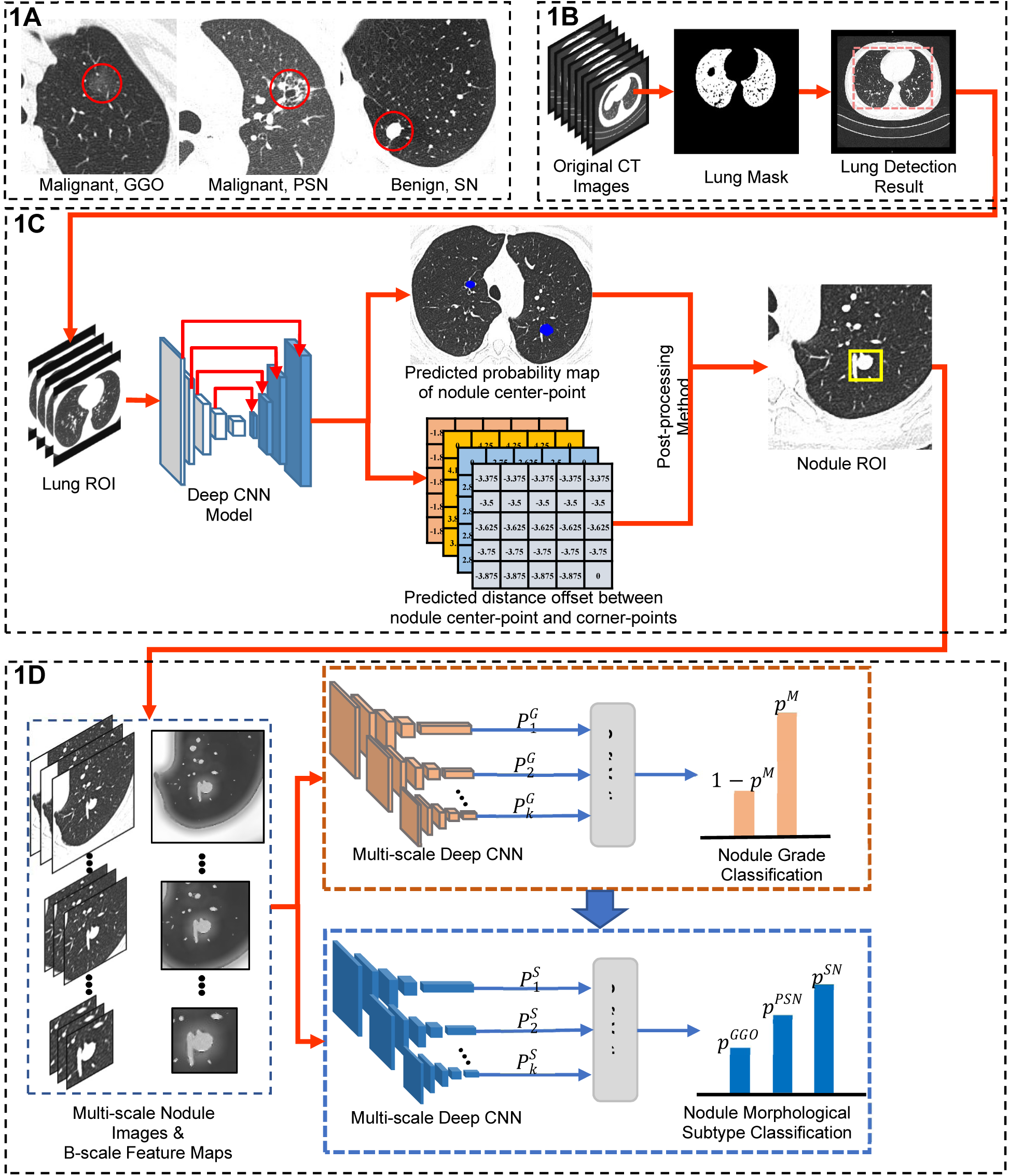
The pipeline of the proposed PN classification system. **(A)** Weak data annotation. The nodule was represented by a red circle. **(B)** Pre-processing method for the CT scans. **(C)** Nodule detection network for automatically locating the PN in the CT scans. **(D)** Nodule classification networks for classifying the grade and subtype labels of nodule.

### Pre-processing for Lung CT Images

The tissue surrounding the lungs produces abundant unnecessary computation. To solve this problem, we designed pre-processing operations including (Figure 1B): i) segmenting the low-intensity regions using simple thresholding for lung tissue which leads to a separation of the of background pixels, and lung object pixels, ii) searching the two regions, excluding the background region and highlighting remaining lung object regions as the lung mask, and finally iii) calculating rectangular boundaries of lung masks and extracting the ROIs of the lung images, which were then resized to 448×448×16 via a linear interpolation and used as the input of deep CNN model for nodule localization.

### Nodule Detection Network

In this study, we designed a center-point (of a nodule) region based object detection network, for which we counted the center of the red circle and the horizontal and vertical distance offset between the top-left and bottom-right points of circle annotation and the center region together as the ground truth labels (Figure 1C). The architecture of the proposed CNN is based on U-Net and VGG-19 (17, 18). The proposed network includes four modules: i) the backbone network (modified VGG-19) for learning multi-scale features from the input image, ii) the feature fusion network for integrating multi-scale feature maps with size 448×448×32, 224×224×32, 112×128×32, 56×56×32, and 28×28×32 together as a feature pyramid, and iii) the pixel-wise classifier for predicting the probability of each pixel attributed to nodule center-point category, and iv) the pixel-wise regressor for predicting the shift between each pixel and top-left and bottom-right points, separately, allowing the predicted distance offset value to be either positive and negative.

To improve the nodule detection performance, we proposed a post-processing method by fusing the center-point and distance offset information. Firstly, the method would search the local-maximum point in center-point pixel-wise classification probability map as the center-point of nodule and compute the coordinate of top-left and bottom-right points of nodule ROI by adding the coordinate of center-point with the offset value. Then, we employed the coordinate of top-left and bottom-right points to extract the nodule ROI from the lung ROI images.

### Nodule Classification Network

We proposed a nodule classification network using the VGG-19 network (18) and the B-scale feature map (Figure 1D), with three steps: i) extracting the multi-scale nodule ROIs from lung ROI, based on the nodule detection result and computing the B-scale feature of the ROI, ii) learning the fusion feature of nodule from the nodule ROI and B-scale feature map, and iii) predicting the grade or type label of nodule ROI by the soft-max classifier. The nodule classification network was built based on VGG-19 network by adding additional the normalization layer and channel attention module into the network to improve the feature learning ability. We also utilized the fully connected layer and soft-max function by predicting the possibility of the input nodule ROI belonging to different nodule grades and sub-type labels. To improve the nodule classification performance, we trained 5 nodule classification networks using 5 different ROI sizes, including 192×192×16, 160×160×16, 128×128×16, 96×96×16, and 64×64×16, and fused the predicted classification possibility by the soft-voting method.

### Model training

We utilized the open source library TensorFlow to implement the proposed nodule detection and classification models and the training data in Table 1 to train models. The hyper-parameters for optimizing the models were as follows: learning rate (0.00001), batch size of training data (50), and number of iterations (500). In addition, our proposed approach is much flexible and allow to utilize different deep learning tools, and including U-Net (17), Nodulenet (19), SANet (20), MSANet (21), AlexNet (22), VGG-19 (18), ResNet-18 (23), and ViT (24). To compare the performance of PN detection methods using different annotations, we utilized the contour, bounding-box, circle, and the point annotations from two datasets to train our model.

## Results

### Evaluation metrics

The precision, recall, and classification accuracy (Acc) were used to quantitatively analyze the nodule detection and classification performance. Besides, we changed the classification thresholding value from 0.05 to 0.95 and computed the true positive rate (TPR) and false positive rate (FPR). In this study, the ROC curve of nodule detection and classification performance is illustrated, in which the horizontal and vertical axis are the Precision and Recall values for nodule detection models and TPR and FPR values for nodule classification models. In addition, we computed the area under the curve (AUC) of ROC as the evaluation metric.

### PN Detection Performance

Our NC-UNet model was compared with other nodule detection models to evaluate their respective performance in nodule detection on LIDC-IDRI and In-house datasets, as shown in Table 2. Our model trained by circle annotations obtained high precision values of 0.929 and 0.936, recall values of 0.932 and 0.959, and AUC values of 0.938 and 0.943 on LIDC-IDRI dataset, which are comparative to the SANet and MSANet models trained by bounding-box annotations. The low standard deviation in these metrics indicates that the proposed approach has good stability and robustness. The Precision-Recall (P-R) curve demonstrated that our model exhibited significantly larger AUC values of 0.932 and 0.964 on LIDC dataset and In-house dataset, respectively, compared to the other three models using circle annotations, indicating our model has a lower false-positive rate and false-negative rate in PN detection (Figure. 2A and 2B).

**Table 2.** Nodule detection performance. Each value is represented as “mean±standard deviation”.

| Annotation | Method | LIDC-IDRI dataset (793 testing cases) |  |  | In-house dataset (822 testing cases) |  |  |
| --- | --- | --- | --- | --- | --- | --- | --- |
|  |  | Precision | Recall | PR-AUC | Precision | Recall | PR-AUC |
| Contour annotation | U-Net (17) | 0.893±0.058 | 0.907±0.041 | 0.915±0.033 | - | - | - |
| Bounding-box annotation | Nodulenet (19) | 0.945±0.049 | 0.965±0.045 | 0.945±0.037 | - | - | - |
|  | SANet (20) | **0.974±0.042** | 0.951±0.053 | 0.937±0.039 | - | - | - |
|  | MSANet (21) | 0.956±0.045 | **0.956±0.037** | **0.952±0.036** | - | - | - |
| Circle annotation | Nodulenet (19) | 0.786±0.085 | 0.824±0.083 | 0.833±0.057 | 0.545±0.089 | 0.775±0.065 | 0.687±0.072 |
|  | SANet (20) | 0.823±0.090 | 0.887±0.084 | 0.862±0.059 | 0.652±0.095 | 0.864±0.084 | 0.726±0.067 |
|  | MSANet (21) | 0.854±0.067 | 0.868±0.058 | 0.875±0.048 | 0.76±0.083 | 0.894±0.068 | 0.804±0.058 |
|  | NC-UNet | 0.929±0.058 | 0.932±0.049 | 0.938±0.046 | **0.936±0.046** | **0.959±0.041** | **0.943±0.035** |
| Point annotation | NC-UNet | 0.855±0.057 | 0.914±0.054 | 0.884±0.065 | 0.837±0.064 | 0.853±0.053 | 0.834±0.042 |

**Table 3.** Nodule grade classification performance. Each value is represented as “mean±standard deviation”.

| Method | LIDC-IDRI dataset |  |  |  | In-house dataset |  |  |  |
| --- | --- | --- | --- | --- | --- | --- | --- | --- |
|  | Acc | TPR | FPR | ROC-AUC | Acc | TPR | FPR | ROC-AUC |
| AlexNet (22) | 0.746±0.064 | 0.694±0.044 | 0.217±0.029 | 0.712±0.063 | 0.635±0.045 | 0.567±0.061 | 0.334±0.056 | 0.672±0.052 |
| VGG-19 (18) | 0.782±0.050 | 0.763±0.032 | 0.124±0.015 | 0.816±0.054 | 0.667±0.050 | 0.612±0.040 | 0.312±0.046 | 0.722±0.077 |
| ResNet-18 (23) | 0.854±0.056 | 0.818±0.039 | 0.113±0.012 | 0.845±0.073 | 0.737±0.072 | 0.635±0.059 | 0.323±0.038 | 0.707±0.046 |
| ViT (24) | 0.819±0.062 | 0.782±0.048 | 0.157±0.018 | 0.798±0.057 | 0.714±0.047 | 0.542±0.035 | 0.267±0.034 | 0.694±0.055 |
| BM-CNN | **0.927±0.039** | **0.898±0.032** | **0.055±0.022** | **0.912±0.037** | **0.768±0.054** | **0.753±0.036** | **0.275±0.029** | **0.815±0.043** |

**Figure 2:**
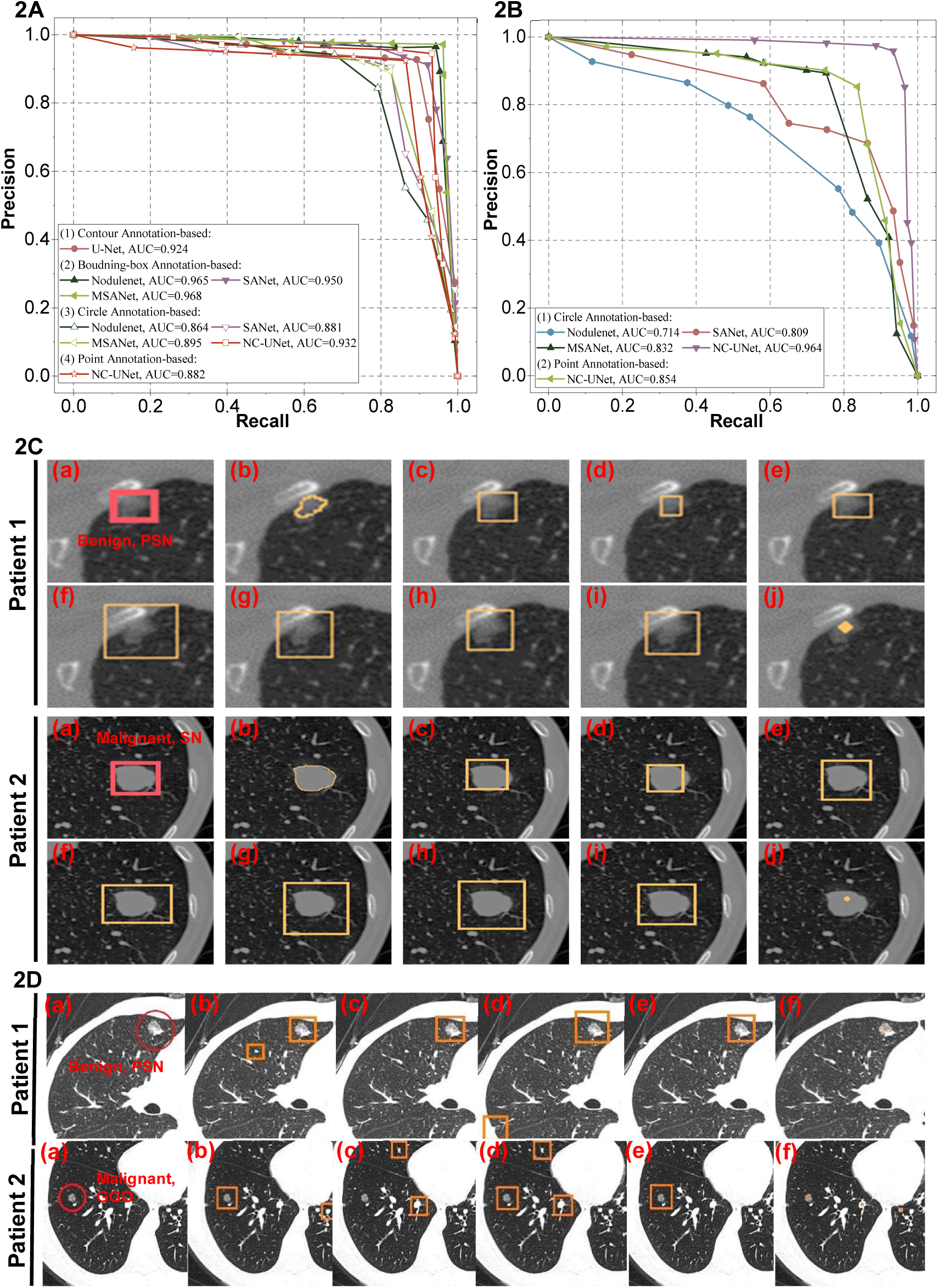
Experimental results of nodule detection. **(A)-(B)** P-R curve of nodule detection performance on LIDC-IDRI and In-house datasets. **(C)** Examples of nodule detection on LIDC-IDRI dataset: (a) bounding-box annotations. (b) nodule segmentation results using U-Net, (c)-(e) nodule detection results using Nodulenet, SANet, and MSANet trained by bounding-box annotations, (f)-(i) nodule detection results using Nodulenet, SANet, MSANet, and NC-UNet trained by circle annotations, (j) nodule detection results using NC-UNet trained by point annotations. **(D)** Examples of nodule detection on private dataset: (a) circle annotations, (b)-(e) nodule detection results using Nodulenet, SANet, MSANet, and NC-UNet trained by circle annotations, (f) nodule detection results using NC-UNet trained by point annotations.

The visualized nodule detection results on LIDC-IDRI and In-house datasets show the excellent agreement between our model and the manual ground truth (Figure. 2C and 2D). By observing the results on LIDC-IDRI dataset, we found that the location and size information of detected nodules for the supervised models trained by bounding-box annotations is more accurate than the models trained by circle annotations, but the difference is not obvious. In addition, the nodule detection results on the In-house dataset demonstrate that our model outperforms other supervised models in terms of nodule detection when using weak annotations to train models.

### PN Classification Performance

We conducted a comparison of the PN grade performance between our BM-CNN model and other models. The ROC curves for nodule classification (Figure. 3A and 3B) on LIDC-IDRI and In-house datasets demonstrate that our model has better nodule classification performance on both two datasets compared with other supervised models. Our model can also predict the morphological subtypes of PN, achieving an overall accuracy of 0.893 and 0.872 on LIDC-IDRI and In-house datasets, respectively. It was the first attempt that hierarchical classification, i.e., nodule grading and subtype classification, was achieved in a single model. The Receiver Operating Characteristic (ROC) curve further revealed that the high accuracy of our model in predicting the morphological subtypes of PN (Figure. 3C and 3D).

**Figure 3:**
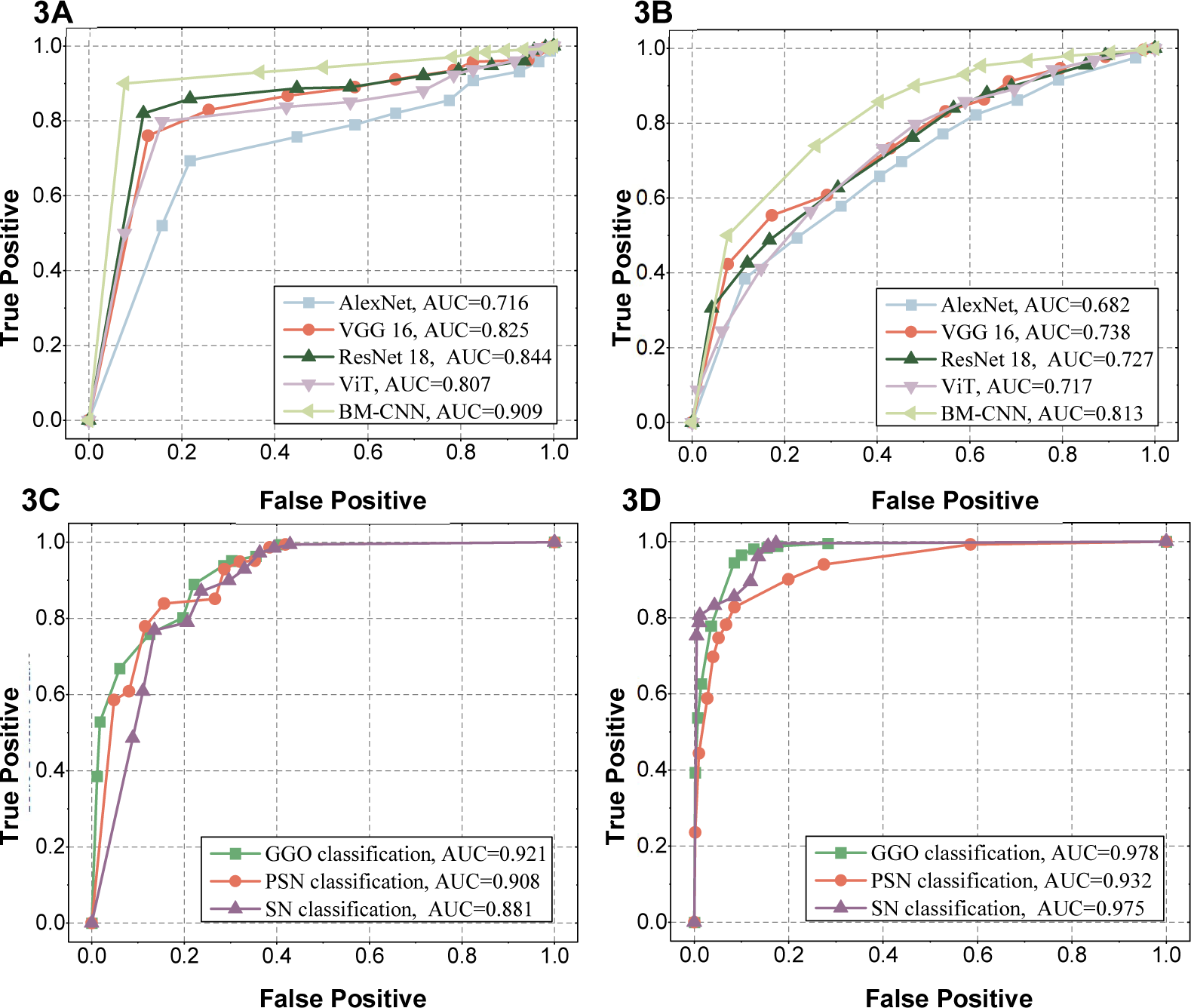
Nodule classification performance of proposed system. **(A)** ROC curve for nodule grade classification on LIDC-IDRI dataset. **(B)** ROC curve for nodule grade classification on In-house dataset. **(C)**. ROC curve for nodule subtype classification on LIDC-IDRI dataset. **(D)** ROC curve for nodule subtype classification on In-house dataset.

As shown in Table 2, our model achieved the highest accuracy values of 0.927 and 0.768 and highest AUC values of 0.912 and 0.815 on LIDC-IDRI and In-house datasets, indicating a low false positive rate and a high true positive rate in nodule grade classification on both two datasets. Moreover, the AUC of 0.912 and 0.815 achieved by our approach is comparable to the state-of-the-arts models, which always used accurate location label and/or nodule contours for their training. The weak annotations in our approach can tolerate more deviations of localization in practical labeling operations, and thus facilitate the robustness of the system (4).

Accordingly, in potential clinical implementation of our model, radiologists will have a significantly reduced burden in labeling the CT images.

Additionally, we conducted an analysis of PN classification performance across nodules with different diameters in our study. Each annotated nodule was measured for its diameter, and we divided the nodules into three subgroups, i.e., small nodules (<10 mm), medium nodules (10-20 mm), and large nodules (20-30 mm). We then used our model to classify the grade (Figure. 4) of nodules in the testing datasets.

**Figure 4:**
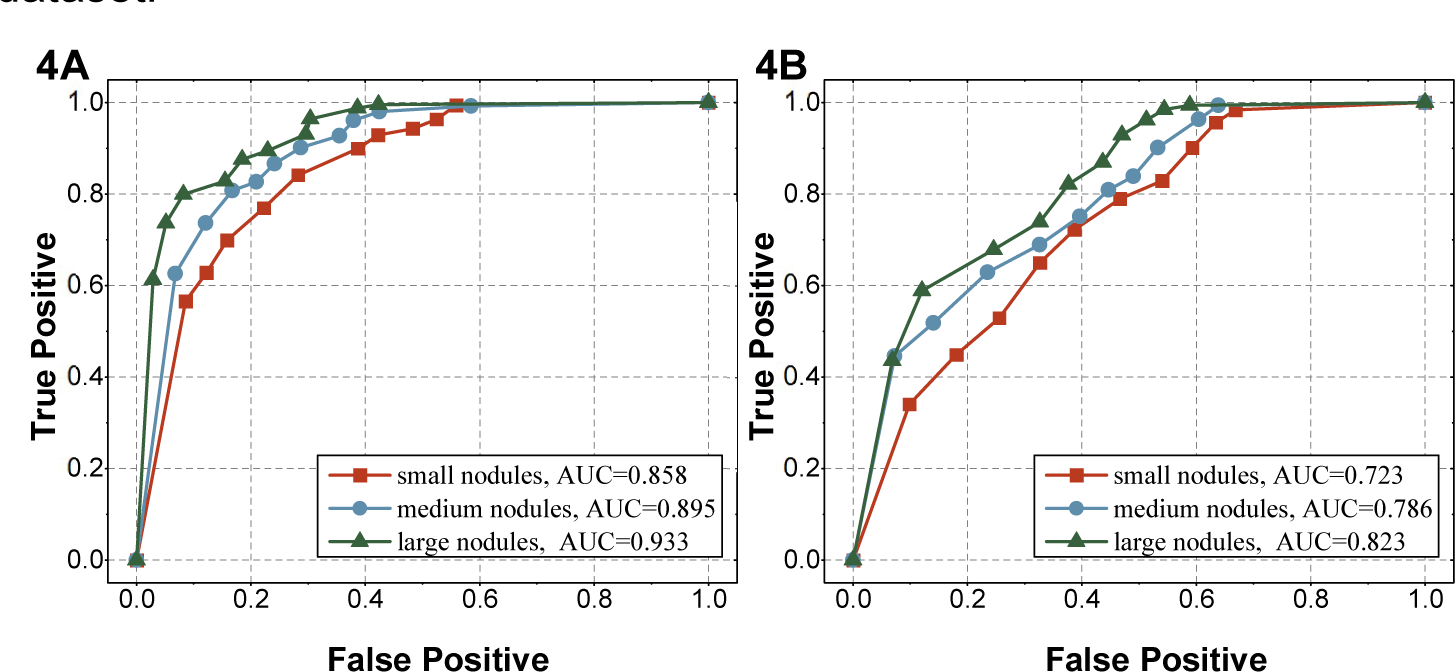
Nodule grade classification performance of proposed system in PN with different diameters. **(A)** ROC curve on LIDC-IDRI dataset. **(B)** ROC curve on In-house dataset.

Our model demonstrated a remarkable grade classification accuracy of 0.879 and 0.712 specifically for small nodules on LIDC-IDRI and In-house datasets, surpassing the average ∼ 30% accuracy of manual nodule classification (25). Furthermore, we observed a consistent improvement in nodule classification accuracy as the nodule diameter increased. This observation suggests that enhancing the clarity of the nodule could enhance nodule classification performance.

### Ablation Study

B-scale as a novel image feature plays a crucial role in establishing a relationship between the segmented training objects and their corresponding images (26). We examined the distribution of B-scale values across different subgroups and conducted statistical analyses to determine the significance (Fig. 5A). We observed a significant divergence in the distribution of B-scale values among the different grade and morphologic subtypes. This finding suggested that the B-scale value of PN in CT images has the potential to serve as a discriminative feature for both PN grade and morphological classification. We conducted a further investigation into the impact of the B-scale feature and soft-voting methods on PN grade classification. (Fig. 5B). We found that both can improve the grade classification performance.

**Figure 5:**
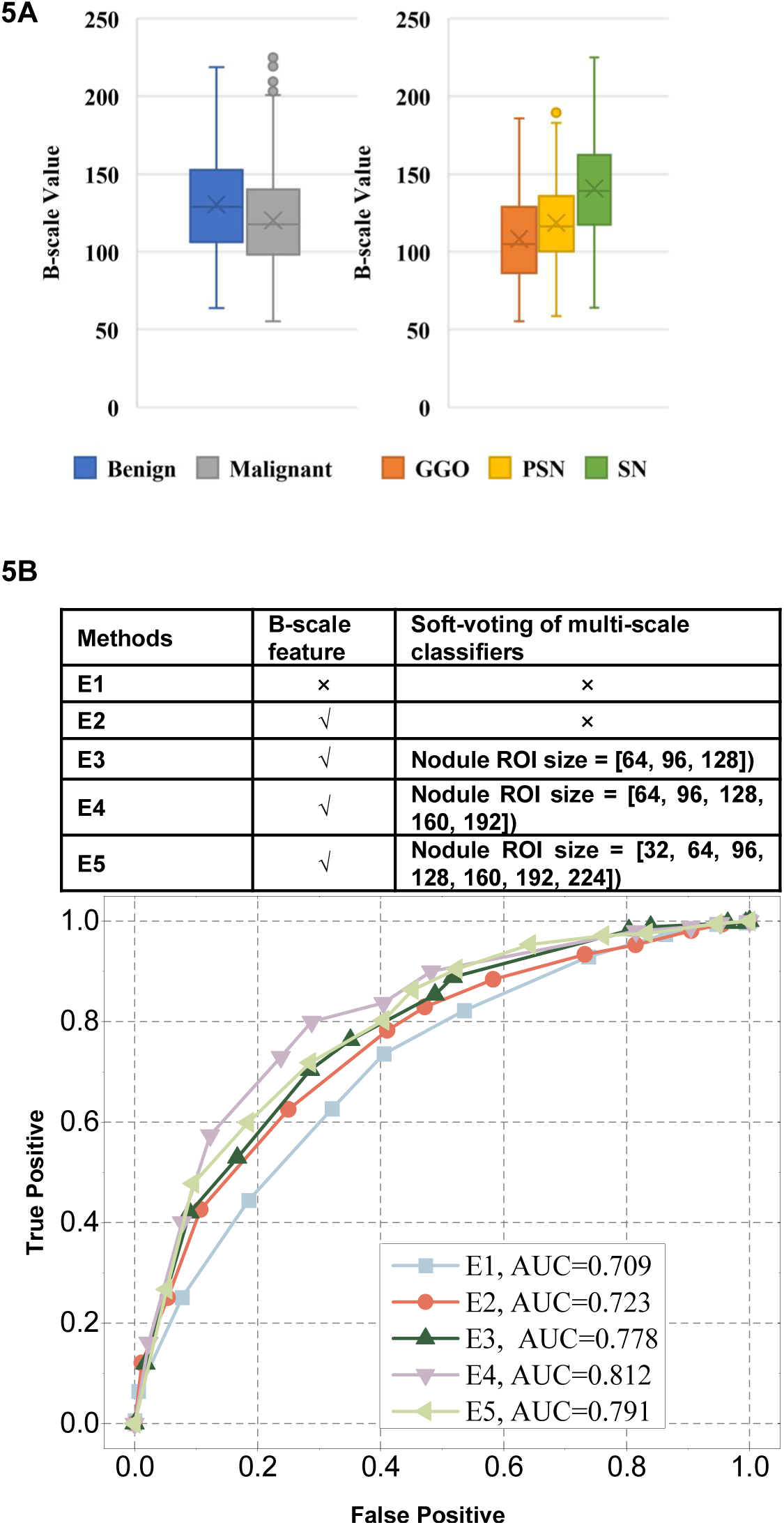
Ablation study on B-scale feature and soft-voting for nodule classification. **(A)** Boxplots for B-scale value distribution in different sub-groups. **(B)** ROC curve of ablation experiments for nodule grade classification.

### Tradeoff Analysis of Model performance and Annotation Time

As shown in Figure 6, we compared four annotation methods regarding annotation time, nodule detection accuracy, and nodule classification accuracy. We utilized the online annotation tool ImgLab^1^ to annotate 10 CT images (including 45 nodules) and time the annotation procedure. Note that the annotation time indicates the total operation time using the annotation tool, excluding the observing time. We found that the PN detection and classification models using circle annotations have approximate accuracy and AUC values with the models using the full annotations, reducing around 30%∼80% of annotation time. In addition, the PN detection and classification models using point annotations have the worst performance due to that the models are unable to learn the nodule size information from the point annotations. Therefore, the circle annotation method has a better tradeoff between data annotation time and PN diagnosis performance.

**Figure 6:**
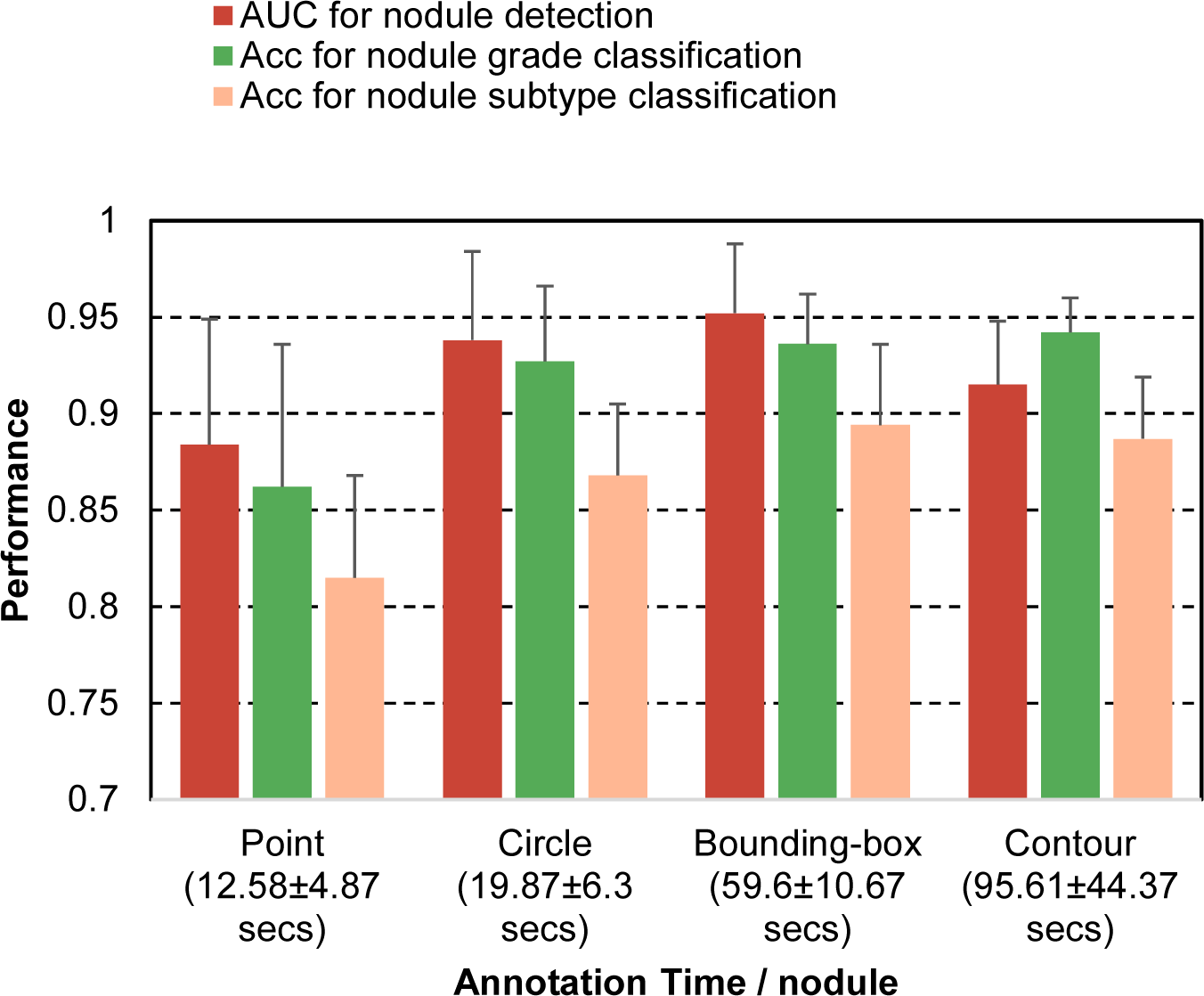
Comparison of the annotation time, PN detection, and classification performance using different annotation methods.

## Discussion

### Data quality of annotation

The majority of present DL approaches rely on extensive and high-quality training datasets (1, 16), which significantly increases the effort amplify the burden of labeling. Medical images pose additional challenges in precise annotation due to the complex anatomical structures, low signal-to-noise ratio, and low contrast among other factors. While using poorly annotated data alleviates these issues, it may produce biased or erroneous models. Therefore, striking a balance between model performances and labeling burden is crucial for an effective nodule classification system. Our weakly supervised PN detection network is capable of accurately locating PN based on coarse annotation results, making our model own higher efficiency and fault tolerance compared to other models that use large and strictly labeled datasets. Therefore, this approach minimizes the time and the cost associated with annotation and enables the construction of an expansive dataset.

These insights derived from this work can also contribute to the development of an efficient system for accurate diagnosis of PN.

### Class-imbalance and image quality in PN classification

We collected and annotated CT images from 2,740 patients with PN and used 822 cases from this dataset to train our model. These dataset sizes are significantly larger than those used in other studies, especially in terms of the testing dataset. As such, our model is more robust compared to other models. Notably, our model showed superior performance in classifying nodule subtypes compared to nodule grade for two primary reasons. The training dataset exhibited an imbalance between MPN and BPN with a ratio of 1:3.56 (Table 4), while the ratio of GGO, PSN, and SN respectively was 1:1.09:1. Compared to other approaches, the class imbalance issue was more challenge in grading nodules in this study. To address the class imbalance concern in nodule grade classification, we used data augmentation techniques to increase the number of BPN and utilized the focal-loss strategy to assign higher weights to the benign label in the loss function. Moreover, we used data in JPEG formatting for the training purpose, which captured the appearance features of the nodules along with compressed tissue attenuation information. The grayscale of the CT images used in this study was only 8-bit, whereas the grayscale of raw CT images typically is 16-bit. This reduction in grayscale resolution by 64-fold can affect the classification of nodule grades. However, the classification of nodule subtypes, which rely on the shape and texture features, would not be impaired by the reduced resolution. Nevertheless, despite these limitations, our model still achieved highly competitive AUC scores for PN grade and subtypes.

**Table 4.**
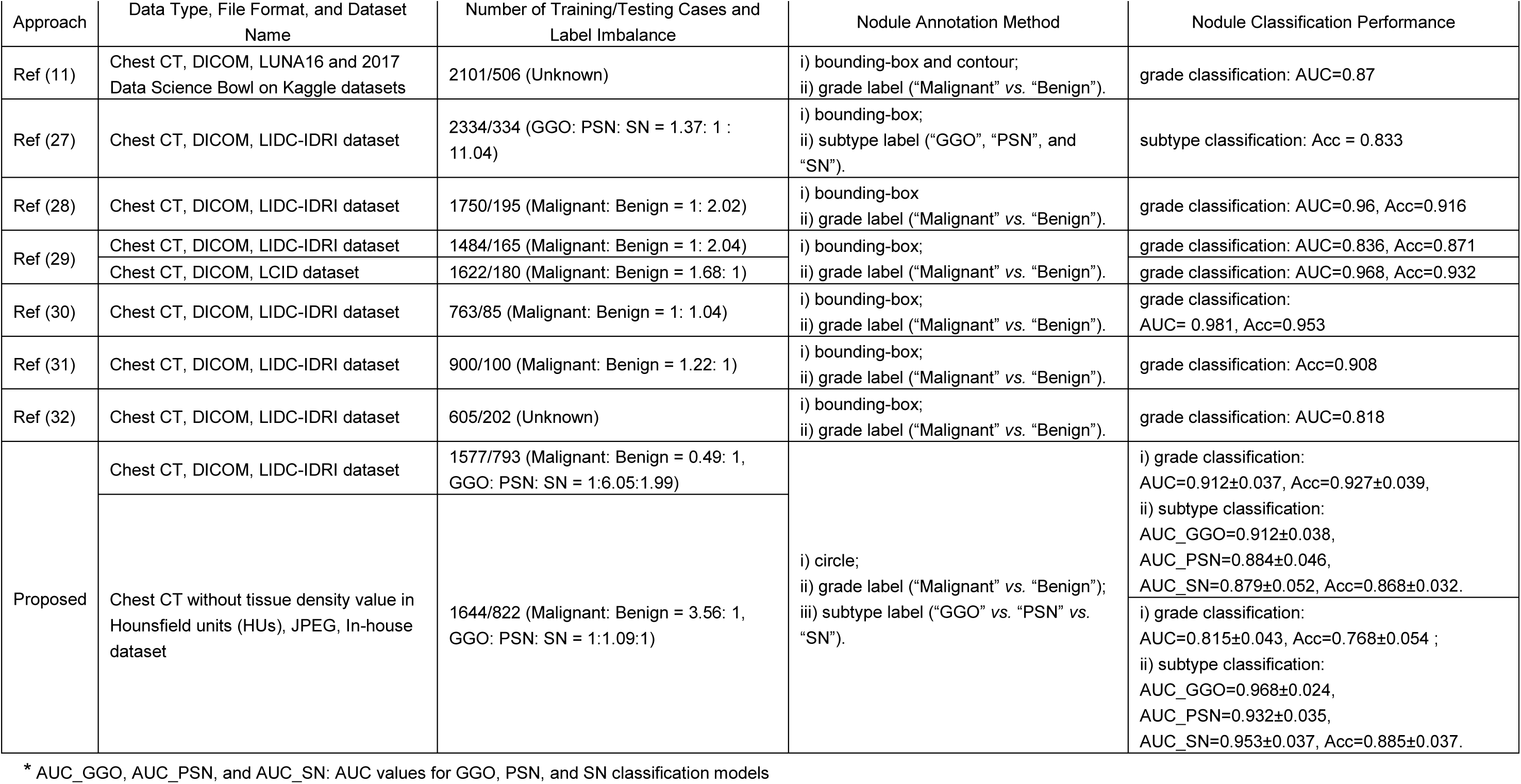
Comparison with other models.

### Handcrafted feature and multi-scale strategy

Our results demonstrated that the incorporation of a handcrafted feature and a multi-scale strategy can significantly enhance the performance of nodule classification. To shed light on this improvement, we used a statistical method to quantitatively assess the variation in B-scale values across different PN grades and subtypes. We found that the use of the B-scale value as prior knowledge guided the model to pay greater attention to shape differences among nodules. In addition, the usage of a multi-scale strategy mitigated the model’s sensitivity to the selection of ROI size, thereby increasing the system’s robustness to variations in nodule size.

Our findings may inspire further research in identifying handcrafted features, which were found to be strongly correlated with nodule classification. New strategies can also be proposed for integrating multi-scale features and classification outcomes, leading to enhanced performance in nodule classification.

### Comparison with the state-of-the-arts

This model encompasses PN detection, PN grade classification, and PN morphologic subtype classification, which closely mimic the manual diagnosis process for PN in clinical settings. Other models have not demonstrated comparable performance thus far. Consequently, our model is better suited for differential diagnosis of PN. Moreover, our model can process CT images in various formats, including DICOM, JEPG, PNG, screen snapshots, and images caught by devices such as those done by cell phone cameras. The versatility allows our model to be implemented in resource-limited settings where different image formats may be encountered. Furthermore, this advantage enhances the speed and safety of medical image transmissions compared to models that solely rely on DICOM files. This breakthrough in the field of medical imaging thus holds tremendous potential for revolutionizing medical data processing and analysis.

## Data Availability

All data produced in the present study are available upon reasonable request to the authors

## Disclosures of conflicts of interest

L.X. No relevant relationships. Y.X. No relevant relationships. M.Z. No relevant relationships. W.M. No relevant relationships.

Y.C. No relevant relationships. M.S. No relevant relationships. M.A.A. No relevant relationships. Y.T. No relevant relationships. Y.W. No relevant relationships.

## Abbreviations

PN: pulmonary nodules
DL: Deep learning
AUC: area under the receiver operating curve
CNN: Convolutional Neural Networks
GGO: pure ground-glass opacity
PSN: part-solid nodules
SN: solid nodules
MPN: malignant PN
BPN: benign PN
ROI: region of interest

## Supplemental Materials

**Figure E1:**
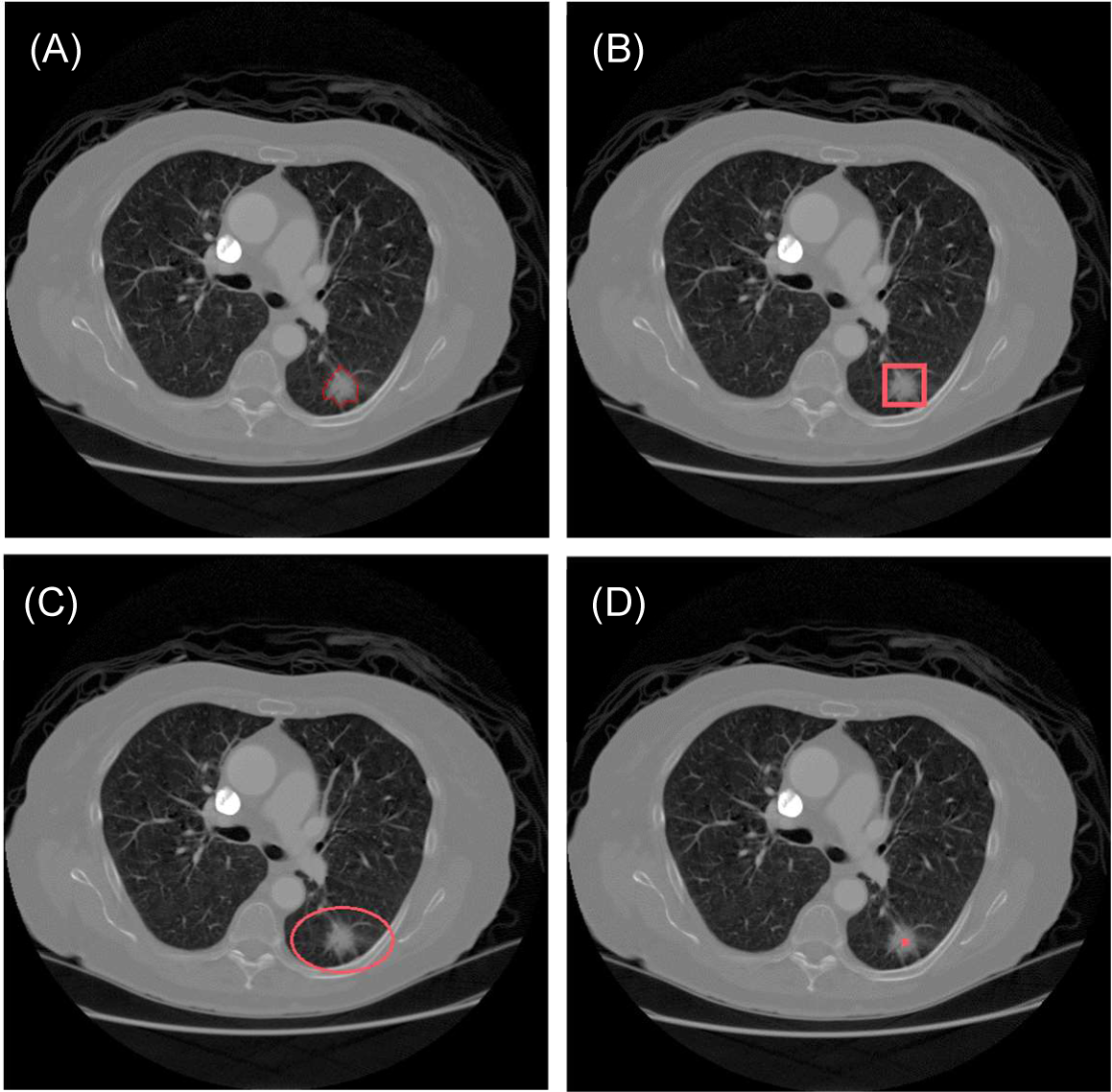
Examples of nodule annotation methods. **(A)** Contour-based annotation. **(B)** Box-based annotation. **(C)** Circle-based annotation. **(D)** Point-based annotation.

**Figure E2:**
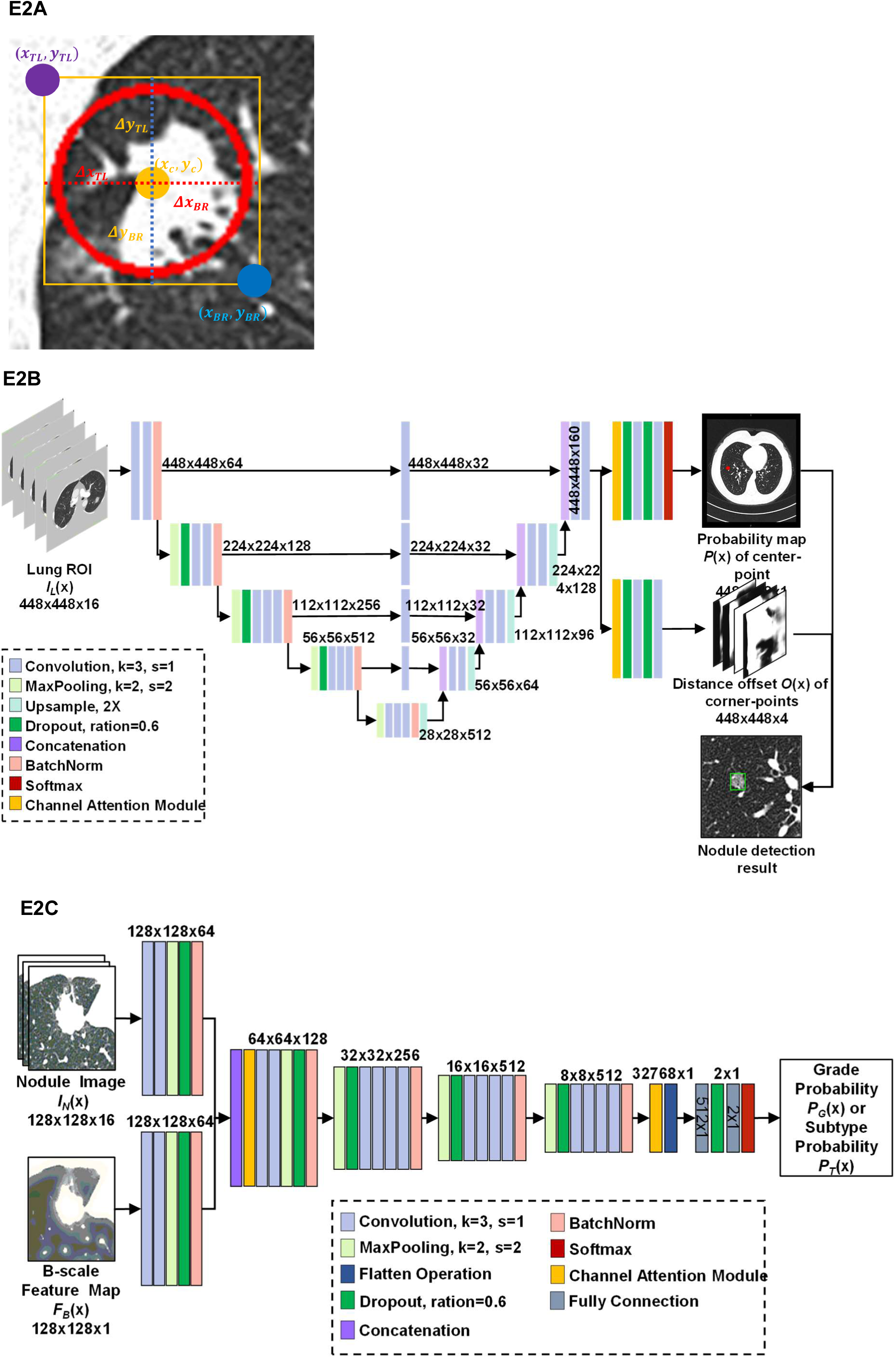
Illustration of PN detection and classification method. **(A)** Example of center, top-right, and bottom-left points of nodule annotation (red circle). The (𝑥_𝑇𝐿_, 𝑦_𝑇𝐿_) and (𝑥_𝐵𝑅_, 𝑦_𝐵𝑅_)represent the location of top-right, and bottom-left points of nodule annotation. The Δ𝑥_𝑇𝐿_, Δ𝑥_𝐵𝑅_, Δ𝑦_𝑇𝐿_, and Δ𝑦_𝐵𝑅_ denote the horizontal and vertical distance offset between the top-left and bottom-right points of circle annotation and the center region. **(B)**The structure of the proposed PN detection network. **(C)** The structure of the proposed PN classification network.

**Figure E3:**
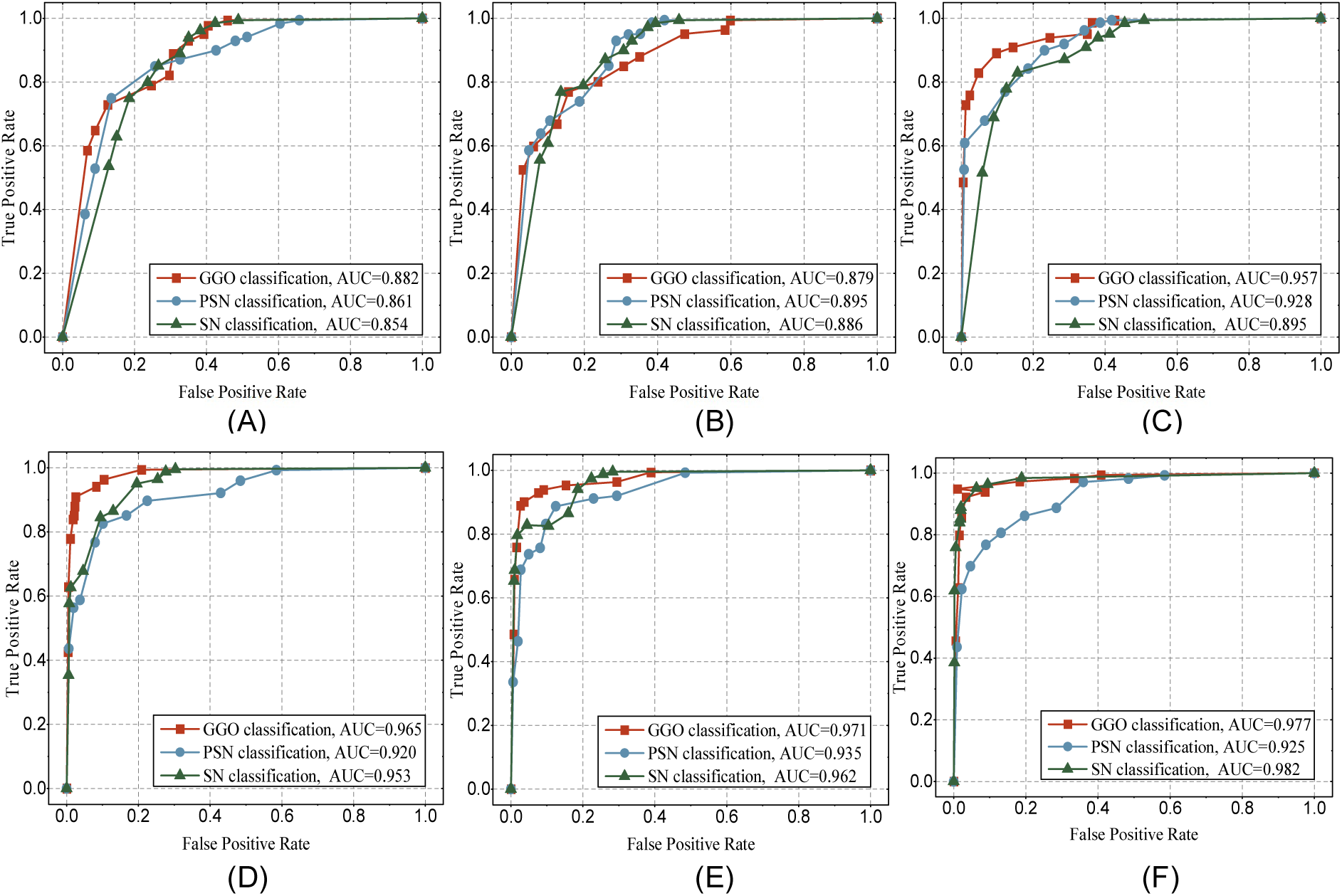
Nodule subtype classification performance of proposed system in PN with different diameters. **(A)-(C)** ROC curves for small, medium, and large nodules on LIDC-IDRI dataset. **(D)-(F)** ROC curves for small, medium, and large nodules on In-house dataset.

**Table E1.**
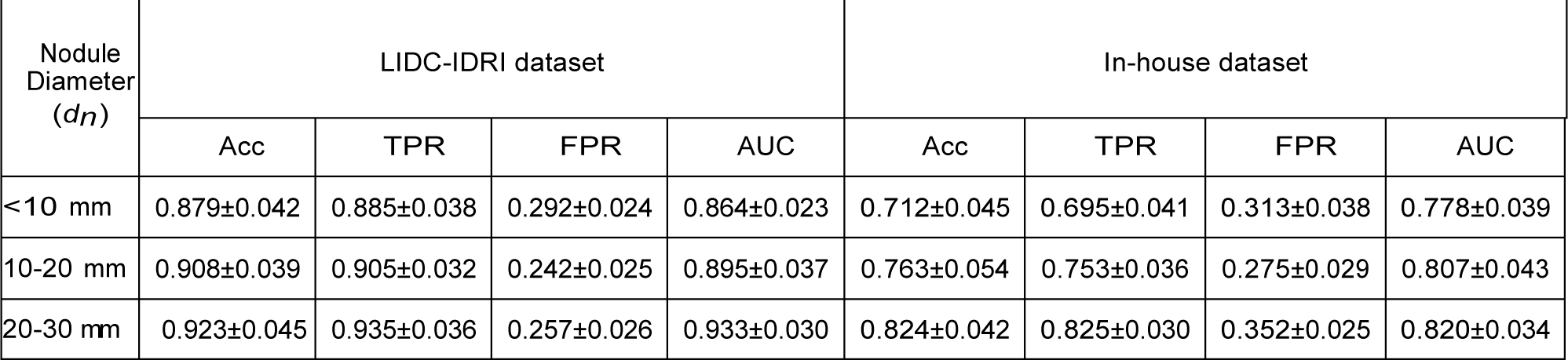
Nodule grade classification performance in nodules with different diameters. Each value is represented as “mean±standard deviation”.

**Table E2.** Nodule subtype classification performance in nodules with different diameters. Each value is represented as “mean±standard deviation”.

| Nodule Diameter ($d_n$) | Nodule Sub-types | LIDC-IDRI dataset | | | | In-house dataset | | | |
| --- | --- | --- | --- | --- | --- | --- | --- | --- | --- |
|  |  | Acc | TPR | FPR | AUC | Acc | TPR | FPR | AUC |
| <10 mm | GGO | 0.842±0.036 | 0.889±0.038 | 0.306±0.024 | 0.882±0.035 | 0.845±0.032 | 0.968±0.015 | 0.085±0.013 | 0.978±0.025 |
|  | PSN | 0.835±0.047 | 0.871±0.054 | 0.326±0.036 | 0.863±0.042 | 0.864±0.037 | 0.862±0.024 | 0.167±0.024 | 0.917±0.034 |
|  | SN | 0.907±0.025 | 0.939±0.047 | 0.352±0.016 | 0.854±0.061 | 0.908±0.048 | 0.887±0.027 | 0.128±0.029 | 0.956±0.042 |
| 10-20 mm | GGO | 0.874±0.045 | 0.879±0.062 | 0.350±0.014 | 0.879±0.072 | 0.905±0.023 | 0.932±0.038 | 0.096±0.016 | 0.968±0.038 |
|  | PSN | 0.866±0.052 | 0.929±0.027 | 0.286±0.017 | 0.857±0.038 | 0.851±0.042 | 0.882±0.034 | 0.163±0.032 | 0.945±0.34 |
|  | SN | 0.903±0.018 | 0.924±0.023 | 0.305±0.021 | 0.886±0.049 | 0.912±0.052 | 0.945±0.042 | 0.045±0.012 | 0.972±0.028 |
| 20-30 mm | GGO | 0.912±0.014 | 0.916±0.017 | 0.143±0.009 | 0.957±0.031 | 0.972±0.027 | 0.987±0.038 | 0.042±0.010 | 0.985±0.016 |
|  | PSN | 0.921±0.024 | 0.929±0.019 | 0.286±0.015 | 0.928±0.042 | 0.863±0.037 | 0.872±0.024 | 0.182±0.024 | 0.928±0.034 |
|  | SN | 0.935±0.029 | 0.938±0.022 | 0.346±0.032 | 0.903±0.017 | 0.917±0.019 | 0.923±0.055 | 0.041±0.009 | 0.985±0.012 |

## Footnotes

1 https://github.com/NaturalIntelligence/imglab/

## References

1. Setio AAA, Traverso A, De Bel T, Berens MS, Van Den Bogaard C, Cerello P, et al. Validation, comparison, and combination of algorithms for automatic detection of pulmonary nodules in computed tomography images: the LUNA16 challenge. Medical image analysis. 2017;42:1–13.

2. Chen K, Bai J, Reuben A, Zhao H, Kang G, Zhang C, et al. Multiomics analysis reveals distinct immunogenomic features of lung cancer with ground-glass opacity. American journal of respiratory and critical care medicine. 2021;204(10):1180–92.

3. Del Ciello A, Franchi P, Contegiacomo A, Cicchetti G, Bonomo L, Larici AR. Missed lung cancer: when, where, and why? Diagnostic and interventional radiology. 2017;23(2):118.

4. Gu Y, Chi J, Liu J, Yang L, Zhang B, Yu D, et al. A survey of computer-aided diagnosis of lung nodules from CT scans using deep learning. Computers in biology and medicine. 2021;137:104806.

5. Gurcan MN, Sahiner B, Petrick N, Chan HP, Kazerooni EA, Cascade PN, et al. Lung nodule detection on thoracic computed tomography images: Preliminary evaluation of a computer─aided diagnosis system. Medical Physics. 2002;29(11):2552–8.

6. Suzuki K, Li F, Sone S, Doi K. Computer-aided diagnostic scheme for distinction between benign and malignant nodules in thoracic low-dose CT by use of massive training artificial neural network. IEEE Transactions on Medical Imaging. 2005;24(9):1138–50.

7. Chen H, Xu Y, Ma Y, Ma B. Neural network ensemble-based computer-aided diagnosis for differentiation of lung nodules on CT images: clinical evaluation. Academic radiology. 2010;17(5):595–602.

8. Zhang F, Song Y, Cai W, Lee M-Z, Zhou Y, Huang H, et al. Lung nodule classification with multilevel patch-based context analysis. IEEE Transactions on Biomedical Engineering. 2013;61(4):1155–66.

9. Kang G, Liu K, Hou B, Zhang N. 3D multi-view convolutional neural networks for lung nodule classification. PloS one. 2017;12(11):e0188290.

10. Ferreira CA, Cunha A, Mendonça AM, Campilho A, editors. Convolutional neural network architectures for texture classification of pulmonary nodules. Iberoamerican congress on pattern recognition; 2018: Springer.

11. Ozdemir O, Russell RL, Berlin AA. A 3D probabilistic deep learning system for detection and diagnosis of lung cancer using low-dose CT scans. IEEE transactions on medical imaging. 2019;39(5):1419–29.

12. Mehta K, Jain A, Mangalagiri J, Menon S, Nguyen P, Chapman DR. Lung nodule classification using biomarkers, volumetric radiomics, and 3D CNNs. Journal of Digital Imaging. 2021:1–20.

13. Tyagi S, Talbar SN. CSE-GAN: A 3D conditional generative adversarial network with concurrent squeeze-and-excitation blocks for lung nodule segmentation. Computers in Biology and Medicine. 2022;147:105781.

14. Khademi S, Heidarian S, Afshar P, Naderkhani F, Oikonomou A, Plataniotis KN, et al., editors. Spatio-Temporal Hybrid Fusion of CAE and SWin Transformers for Lung Cancer Malignancy Prediction. ICASSP 2023-2023 IEEE International Conference on Acoustics, Speech and Signal Processing (ICASSP); 2023: IEEE.

15. Mkindu H, Wu L, Zhao Y. Lung nodule detection in chest CT images based on vision transformer network with Bayesian optimization. Biomedical Signal Processing and Control. 2023;85:104866.

16. Armato III SG, McLennan G, Bidaut L, McNitt-Gray MF, Meyer CR, Reeves AP, et al. The lung image database consortium (LIDC) and image database resource initiative (IDRI): a completed reference database of lung nodules on CT scans. Medical physics. 2011;38(2):915–31.

17. Ronneberger O, Fischer P, Brox T, editors. U-net: Convolutional networks for biomedical image segmentation. International Conference on Medical image computing and computer-assisted intervention; 2015: Springer.

18. Simonyan K, Zisserman A, editors. Very deep convolutional networks for large-scale image recognition. 3rd International Conference on Learning Representations (ICLR 2015); 2015: Computational and Biological Learning Society.

19. Tang H, Zhang C, Xie X, editors. Nodulenet: Decoupled false positive reduction for pulmonary nodule detection and segmentation. Medical Image Computing and Computer Assisted Intervention 2019, Proceedings, Part VI 22; 2019: Springer.

20. Mei J, Cheng M-M, Xu G, Wan L-R, Zhang H. SANet: A slice-aware network for pulmonary nodule detection. IEEE transactions on pattern analysis and machine intelligence. 2021;44(8):4374–87.

21. Guo Z, Zhao L, Yuan J, Yu H. Msanet: multiscale aggregation network integrating spatial and channel information for lung nodule detection. IEEE Journal of Biomedical and Health Informatics. 2021;26(6):2547–58.

22. Krizhevsky A, Sutskever I, Hinton GE. Imagenet classification with deep convolutional neural networks. Communications of the ACM. 2017;60(6):84–90.

23. He K, Zhang X, Ren S, Sun J, editors. Deep residual learning for image recognition. Proceedings of the IEEE conference on computer vision and pattern recognition; 2016.

24. Dosovitskiy A, Beyer L, Kolesnikov A, Weissenborn D, Zhai X, Unterthiner T, et al., editors. An Image is Worth 16×16 Words: Transformers for Image Recognition at Scale. International Conference on Learning Representations; 2020.

25. Saji H, Okada M, Tsuboi M, Nakajima R, Suzuki K, Aokage K, et al. Segmentectomy versus lobectomy in small-sized peripheral non-small-cell lung cancer (JCOG0802/WJOG4607L): a multicentre, open-label, phase 3, randomised, controlled, non-inferiority trial. The Lancet. 2022;399(10335):1607–17.

26. Bagci U, Udupa JK, Chen X, editors. Ball-scale based hierarchical multi-object recognition in 3D medical images. Medical Imaging 2010: Image Processing; 2010: SPIE.

27. Ferreira CA, Cunha A, Mendonça AM, Campilho A, editors. Convolutional Neural Network Architectures for Texture Classification of Pulmonary Nodules 2019; Cham: Springer International Publishing.

28. Xie Y, Xia Y, Zhang J, Song Y, Feng D, Fulham M, et al. Knowledge-based collaborative deep learning for benign-malignant lung nodule classification on chest CT. IEEE transactions on medical imaging. 2018;38(4):991–1004.

29. Fu Y, Xue P, Xiao T, Zhang Z, Zhang Y, Dong E. Semi-Supervised Adversarial Learning for Improving the Diagnosis of Pulmonary Nodules. IEEE Journal of Biomedical and Health Informatics. 2022.

30. Al-Shabi M, Shak K, Tan M. ProCAN: Progressive growing channel attentive non-local network for lung nodule classification. Pattern Recognition. 2022;122:108309.

31. Jiang H, Shen F, Gao F, Han W. Learning efficient, explainable and discriminative representations for pulmonary nodules classification. Pattern Recognition. 2021;113:107825.

32. Harsono IW, Liawatimena S, Cenggoro TW. Lung nodule detection and classification from Thorax CT-scan using RetinaNet with transfer learning. Journal of King Saud University-Computer and Information Sciences. 2022;34(3):567–77.

